# Continuous Value Tokenization Improves Medical Event Foundation Models

**DOI:** 10.64898/2026.08.04.26359713

**Authors:** Kent A McCann, Ikgyu Shin, Huan Li, Davis White, Edward R Melnick, Mark Iscoe, Andrew J Loza

**Affiliations:** Department of Emergency Medicine, Yale School of Medicine, New Haven, CT; Department of Biomedical Informatics and Data Science, Yale School of Medicine, New Haven, CT; Epic Systems; Yale New Haven Health System, New Haven, CT

## Abstract

Medical foundation models convert patient records into token sequences for autoregressive prediction, but numeric values such as lab results, vital signs, and time intervals are typically discretized into bins, losing precision and misaligning with clinical thresholds. We trained decoder-only transformer models (47 million parameters) on MIMIC-IV data (364,627 patients; 375 million observations) to compare three tokenization strategies: Discrete (binned values), Continuous Factored (continuous values preserving sequence length), and Continuous Fused (continuous values fused with measurement-type tokens). We evaluated next-token prediction, numeric value prediction, and three clinical tasks: ED disposition at triage, ICD code prediction, and DRG prediction at discharge. Continuous Fused tokenization reduced median sequence length by 34%, reached the Discrete model’s final next-token loss in 30% of training iterations, and improved numeric prediction accuracy by 30.25% median nRMSE reduction. ICD code prediction favored Continuous Fused (AU-PRC 0.457 vs. 0.446; p < 0.001); DRG prediction was equivalent between Continuous Fused and Discrete; ED disposition accuracy was equivalent across all models (∼0.900), though Discrete achieved better calibration. We additionally explain why predictive performance improves with Monte Carlo sample count and derive a scaling law to predict performance gains from increasing simulation budget. Continuous-value tokenization offers substantial efficiency and precision gains while maintaining comparable clinical task performance, with no modifications to the standard transformer architecture.

**Author Summary:** Medical event foundation models are increasingly used to forecast clinical outcomes. These models predict using patient records as sequences of “tokens,” much like a language model predicts next words. Numerical values such as lab results and vital signs pose a problem: they are usually grouped into discrete bins before being tokenized, which loses precision and ignores the fact that medical decisions often depend on specific numerical thresholds. We compared three ways of representing continuous values in medical foundation models: the standard binning approach and two new approaches that preserve the actual values. The continuous-value approaches trained about three times faster, produced 34% shorter sequences, and achieved comparable accuracy on clinical prediction tasks including emergency department triage, ICD diagnosis codes, and hospital billing codes. These gains require no modifications to the standard transformer architecture. We also discovered that commonly used evaluation metrics exhibit a systematic bias depending on the number of simulated predictions, and that this bias follows a predictable mathematical pattern, enabling researchers to estimate full-scale performance from smaller, less expensive simulation experiments.

## Introduction

Foundation models have been proposed as a general-purpose paradigm for artificial intelligence in medicine [1,2]. Recent medical event foundation models generate predictions across clinical event risk, diagnosis, disease progression, and patient flow, as exemplified by Curiosity [3], ETHOS [4], Delphi [5], MOTOR [6], and CLMBR [7]. These models leverage the same transformer architecture as large language models [8], except that instead of modeling sequences of words or subwords, they model the sequence of medical events. This requires converting the data in a patient’s longitudinal medical record into a sequence of tokens, a process that presents a particular challenge for numeric quantities such as lab values, vital signs, and time. Many current medical event foundation models handle numeric clinical quantities through discretization, converting continuous values to discrete bins that represent a range of numeric values and assigning each bin a discrete token [3,4,9]. Related EHR transformer models similarly rely on finite categorical event vocabularies rather than continuous numeric representations [5,10]. For numeric observations, discretization leads to two significant and related limitations. First, discretization causes information loss, preventing models from leveraging numeric differences among observations within the same bin. Second, discretization results in misalignment with diagnostic and treatment thresholds, as many medical decisions and diagnostic definitions rely on specific numerical thresholds that may fall within a given bin [11].

To retain precision and align with necessary downstream use cases, medical event foundation models may benefit from a continuous representation of numeric values. Architectures like Event Stream GPT [12] and HVAT [13] achieve high fidelity by explicitly embedding continuous values, but they introduce model complexity that departs from a standard autoregressive transform design. In prior work, we developed multivariateGPT, a method for integrating text, categorical, and numeric data into generative pretrained transformer training [14]. This approach has two advantages over existing methods: it preserves the standard language model architecture, and it reduces sequence length compared to the tokenization used in ETHOS and Curiosity. While this method demonstrated improved performance on small-scale Electronic Health Record (EHR) tasks, it has not yet been scaled to a full, large-scale EHR dataset.

In this study, we test whether this continuous-value tokenization procedure improves the performance of a full-scale medical event foundation model. We compare it with a discrete tokenization baseline similar to that used in ETHOS and Curiosity [3,4]. This work makes four significant contributions. First, we demonstrate that continuous-value representation achieves comparable clinical prediction performance while substantially reducing training time and sequence length. Second, we show that this tokenization approach can reduce the training iterations required to match the discrete baseline’s final loss by 70%. Third, we present a pipeline that integrates directly into the Medical Event Data Standard (MEDS) ecosystem and works with any generative pretrained transformer backbone with minimal modifications. Finally, we provide an explanation for the previously observed phenomenon that predictive performance improves with Monte Carlo sample count, and we derive a scaling law that allows researchers to predict performance gains from increasing simulation budget.

## Methods

### Ethics Statement

This study used the Medical Information Mart for Intensive Care (MIMIC-IV) dataset (versions 3.1 for the *core*, *hosp*, and *icu* domains; version 2.2 for the *ed* domain), a de-identified electronic health record dataset of patients cared for at Beth Israel Deaconess Medical Center (BIDMC) between 2008 and 2022 [15,16]. MIMIC-IV was created under approval from the BIDMC Institutional Review Board with a waiver of informed consent, and data were de-identified in compliance with the HIPAA Safe Harbor provision. Access is governed by the PhysioNet credentialed-access Data Use Agreement (DUA), which requires completion of human-subjects research training and signed acknowledgment of data use restrictions. All authors completed the required training and agreed to the PhysioNet DUA prior to data access. This study was reviewed by the Yale Institutional Review Board and determined to be exempt under 45 CFR 46.104(d)(4). Patients and the public were not involved in the design, conduct, or reporting of this study, as it relies on a pre-existing de-identified administrative dataset and addresses a methodological question in model development. The work conforms to the principles of the Declaration of Helsinki [17] and the Belmont Report [18].

### Dataset

Models were trained on the MIMIC-IV dataset described above, which contains de-identified health data for more than 300,000 patients. Data were extracted using the MIMIC-IV-MEDS repository [19] and converted to the MEDS format [20]. Every row in the MEDS-formatted dataset represents a medical observation. These observations can be either categorical or numeric. We included the 200 most frequently observed numeric observations in our dataset, as done in ETHOS [4]. We excluded a limited number of observations with data quality issues that included duplicate ED stays and temporally invalid events (78 of approximately 425,000 ED visits). Numeric observations for which no value was recorded were omitted from the sequence entirely; no imputation was performed. The data were split into the following sets: training (80%), validation (10%), and test (10%). These same sets were used for all model training and evaluation throughout the study.

### Tokenization

Tokenization closely followed the methods described in ETHOS [4], however instead of using custom time-delta token bins to capture the passage of time, we defined two time-delta token types, one for intervals less than 24 hours and one for intervals greater than 24 hours, with elapsed time represented continuously. A table of tokens derived from each class of medical data (e.g., diagnoses, medications) is provided in **S1 Table**.

We compared three tokenization methods for numeric values, as shown in **Figure 1** and explained in detail in the following sections. To ensure fair comparison between approaches, the categorical observations in the dataset for each model were identical. For each tokenization method, we report an overall vocabulary size and breakdown by category as well as patient record sequence lengths.

**Figure 1:**
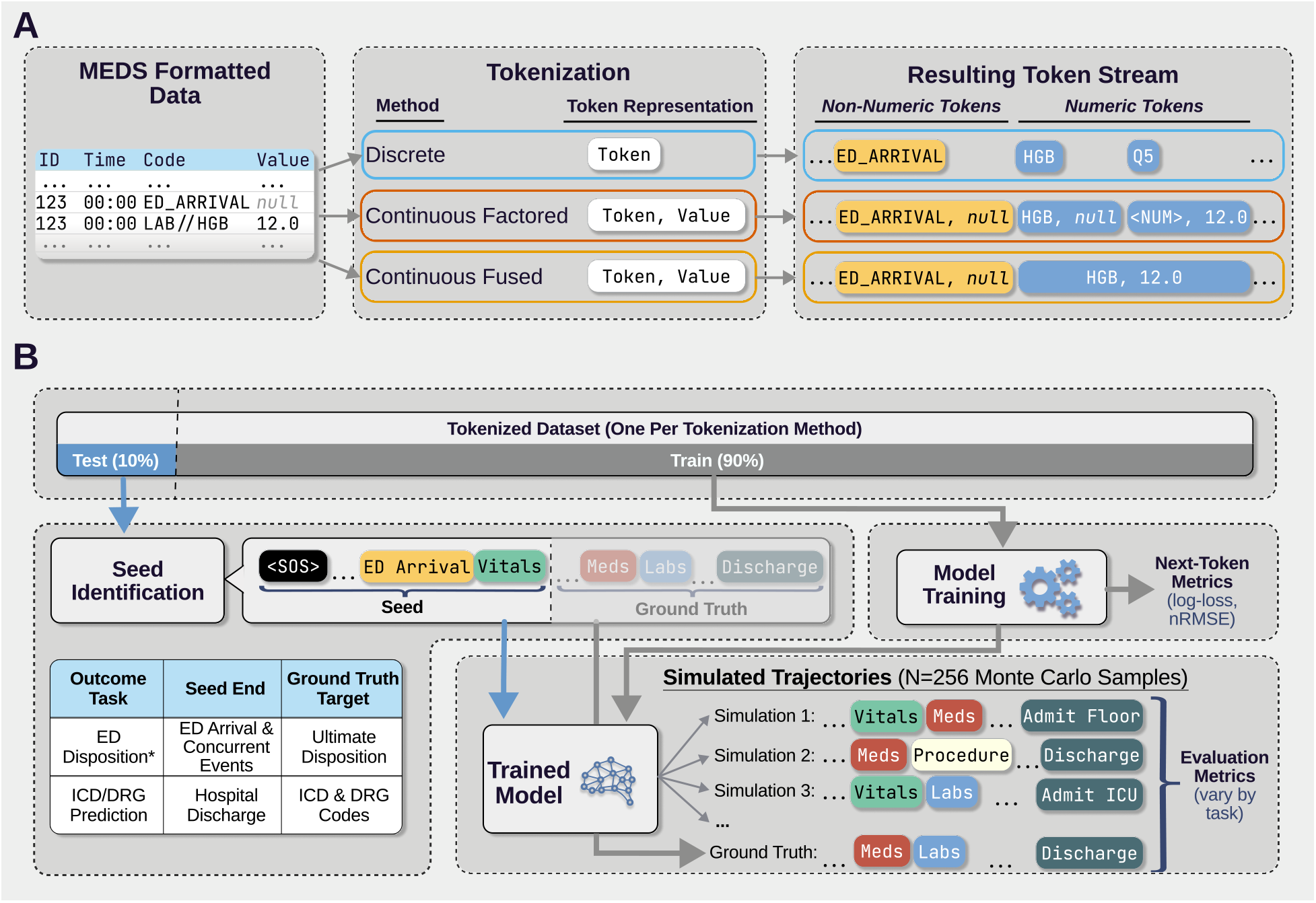
Overview of tokenization methods and data pipeline. (A) Two simultaneous MEDS events, a categorical event (ED_ARRIVAL) and a numerical event (LAB//HGB), and the resulting token stream for each tokenization method. In Continuous representations, all observations are encoded as (Token, Value) tuples, with non-numeric observations carrying a null value. In Continuous Factored, the value is associated with a separate ⟨NUM⟩ token; in Continuous Fused, it is associated directly with the measurement-type token. (B) Models were evaluated on next-token accuracy metrics (training loss and nRMSE) and three clinical prediction tasks: ED disposition at triage and ICD and DRG codes at hospital discharge. For each task, patient sequences were truncated at the prediction point to create “seed” contexts (2,000 for ED disposition; 1,999 for ICD code prediction; 1,646 for DRG-HCFA; 1,525 for DRG-APR). 256 Monte Carlo samples were generated per seed and aggregated against ground-truth outcomes. *Seed identification and Monte Carlo sampling for the ED Disposition task are shown in this example*.

### Numeric Tokenization Methods

Numeric observations include a token identifying the measurement type (e.g., creatinine, heart rate) and an associated numeric value. We kept the measurement-type token constant between approaches and only varied how the accompanying numeric value was represented. We compared three numerical value representation strategies that we refer to as 1) Discrete, 2) Continuous Factored and 3) Continuous Fused. Example token sequences for each are shown in **Figure 1**A.

### Discrete Value Representation

In the Discrete approach, values for each numeric observation were mapped to one of 10 equal-width bins spanning the 1st–99th percentile range in the training set. Values outside this range were truncated to the boundary bins. The resulting bin index was encoded as a separate token that immediately followed the measurement-type token (Figure 1A), consistent with the approach used in ETHOS and Curiosity [3,4].

### Continuous Value Representation

In continuous representations, numeric values were retained as continuous scalar values. For each measurement type, values were first truncated to the training-set 1st–99th percentile range. Then values were normalized by fitting each numeric observation class to a set of candidate distributions (z-score, log-normal, gamma, or min-max) and selecting the distribution that minimized the deviation between the transformed data’s quantiles and the theoretical z-scores of a standard normal distribution. Raw numeric values were then transformed using the selected distribution’s cumulative density function. The resulting normalized scalar was provided to the model as a continuous value associated with the corresponding measurement-type token. As a result, all observations in Continuous approaches are encoded as tuples of (Token, Value), with non-numeric observations carrying a null value.

We evaluated two methods for including the continuous value within the token stream. In the Continuous Factored representation, each numeric observation was represented by the measurement-type token followed by a dedicated numeric concept token <NUM>. The continuous value was associated with <NUM>, mirroring the two-token structure of the discrete baseline while changing only the value representation. In the Continuous Fused representation, the continuous value embedding was attached directly to the measurement-type token, eliminating the additional <NUM> token and thereby reducing sequence length. We evaluated both to help distinguish the effects of continuous value representation from the effects of sequence-length reduction (Figure 1A).

### Model Training

The models were implemented as decoder-only transformers following nanoGPT-style architecture [21]. All tokenization schemes leveraged the same core model architecture: 10 attention heads and 10 transformer layers with an embedding dimension of 600, yielding approximately 47 million trainable parameters. Training was conducted with a context window of 512 tokens and a total batch size of 131,072 tokens. All models were trained on approximately 6.6 billion tokens for 50,000 iterations using the AdamW optimizer with a learning rate of 0.0015 (selected as optimal for the Discrete model), weight decay of 0.05, and momentum parameters *β*_1_ = 0.9 and *β*_2_ = 0.95. Hyperparameters were selected based on established defaults for autoregressive language models of this scale, with minor adjustments during preliminary experiments on validation loss. All experiments were conducted on a single NVIDIA H200 GPU.

### Next-Token Performance Comparison

To ensure fair loss comparison, we computed a masked categorical token-only cross-entropy loss (masked validation loss): the cross-entropy loss evaluated only at positions where the model predicts categorical-type tokens, and excluded numeric-type tokens. To evaluate numeric prediction accuracy, we computed the class normalized Root Mean Square Error (nRMSE) for each numeric observation across 50 consecutive test batches. The RMSE was computed for each numeric observation class and normalized by dividing by the mean of each numeric class. Error was computed on predicted values after reversing the discretization (discrete) or centering and scaling (continuous). For the discrete model, we converted the predicted bin to a numeric value by mapping it to the midpoint of the corresponding bin. For the continuous model, we applied the inverse of the observation-specific normalization transform to the predicted scaled value.

### Clinical Outcome Prediction

We performed three predictive tasks for each model: predicting ED disposition at the time of triage and predicting ICD and DRG codes at the time of hospital discharge. For each task, we identified relevant patient sequences up to the point at which the simulated token sequence was to begin. We referred to this as the seed. The prediction instant, defined as the timepoint at which the model transitions from observed to simulated data, is the time of triage for ED disposition and the time of hospital discharge for ICD and DRG tasks; the seed (i.e., the model prompt) comprises the patient’s event sequence up to the prediction instant. For ED disposition, we defined the seed end as the last event with the same time stamp as the time of arrival to the ED. For ICD and DRG code prediction, we identified the seed end as the hospital discharge event token. We always used the beginning of the patient’s medical event sequence (the <SOS> token, which serves as a beginning-of-sequence marker for each patient) as the seed start. A representation of this process is shown in **Figure 1**B.

We identified two evaluation cohorts from the test set: 2,000 ED encounters for the disposition task and 2,000 hospital discharges for the ICD and DRG tasks. The same hospital discharge cohort was used for both ICD and DRG prediction. If the seed exceeded the context window, we retained the maximum number of recent tokens while still allowing for generation of the next token. Monte Carlo sampling was used to estimate clinical outcomes by generating multiple predicted future sequences for each seed and computing the type and frequency of each outcome. We performed 256 Monte Carlo (MC) samples per seed, yielding 512 tokens for each simulation. Simulations that did not produce a valid endpoint token within the 512-token generation window were included in the analysis with their partial event sequences. Simulations were performed in parallel batches on H200 GPUs hosted on an institutional compute cluster. No class-imbalance correction was applied during either training or evaluation; autoregressive next-token prediction treats all token positions uniformly, and class frequency stratification in the evaluation (Table 4) serves as the reporting strategy for imbalanced outcome distributions. Task-specific evaluation methods were as follows:

For ED Disposition, we computed the frequency of each disposition class across MC samples. The potential dispositions were Admit Hospital, Admit ICU, Transfer, Home Intended, Home Unintended, Other, and Expired. Home Unintended included elopement and discharge against medical advice; Other encompassed observation and other non-standard dispositions. Performance was summarized using top-1 accuracy. Calibration was assessed using calibration curves and Expected Calibration Error (ECE), computed using 10 uniform-width probability bins.

For ICD code prediction, we evaluated discriminative performance using precision-recall curves generated by sweeping inclusion thresholds across the full range of MC code frequencies (1/256 to 255/256). Overall discriminative performance was summarized as the area under the precision-recall curve (AU-PRC), comparing AU-PRC as a function of MC sample count.

For DRG code prediction, we analyzed predictive ability on two classification systems: the Health Care Financing Administration (HCFA) system and the All Patient Refined (APR) system. Each encounter contained one DRG code per system, and we evaluated them as two separate single-label classification tasks. For each system, we generated PR curves by sweeping inclusion thresholds on MC code frequencies, identical to the ICD approach.

To evaluate performance across code frequencies, we stratified ICD and DRG codes into three prevalence bins based on their frequency within the test set. Bin boundaries were selected such that common, moderate, and rare codes account for approximately 75%, 20%, and 5% of total code occurrences, respectively. AU-PRC was computed and compared for each model within these frequency bands.

### Monte Carlo Sample-Count Sensitivity Analysis

To characterize how measured performance changes with the number of Monte Carlo simulations, we recomputed all clinical-task evaluation metrics at *N* = 4, 8, 16, 32, 64, 128, 256 samples. For each metric and tokenization variant, we fitted a scaling law to the observed prefix trajectory through a maximum of half the target sample count, then extrapolated to predict performance at the target *N*. Three prediction horizons were evaluated: predicting *N* = 64 from *N* ≤ 32, *N* = 128 from *N* ≤ 64, and *N* = 256 from *N* ≤ 128. Performance was assessed as the absolute error between the predicted and observed metric value at the target sample count.

## Results

### Tokenization, Vocabulary, and Resulting Dataset

The population of the MIMIC-IV database is described in detail in [15] and [16]. The dataset comprises 375,419,756 total observations from 364,627 unique patients. **Table 1** summarizes the model vocabulary for each tokenization strategy, and **Table 2** reports the resulting per-patient sequence length distributions. Continuous Fused tokenization yielded median sequence lengths 34% shorter than those of factored approaches, although both exhibited heavy-tailed distributions, with the longest patient timeline exceeding 470,000 tokens under Discrete tokenization.

**Table 1:**
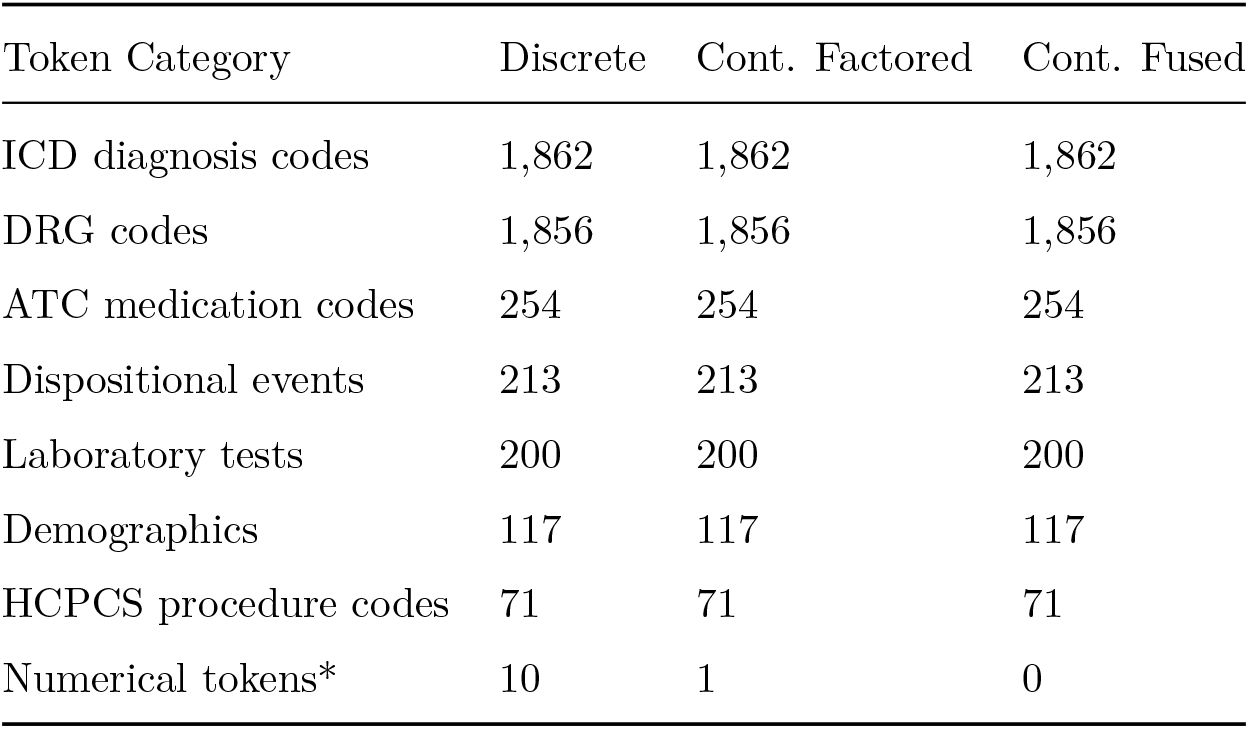
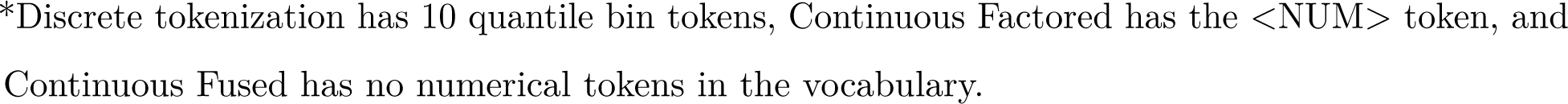
Model vocabulary by token category and tokenization strategy.

| Token Category | Discrete | Cont. Factored | Cont. Fused |
| --- | --- | --- | --- |
| ICD diagnosis codes | 1,862 | 1,862 | 1,862 |
| DRG codes | 1,856 | 1,856 | 1,856 |
| ATC medication codes | 254 | 254 | 254 |
| Dispositional events | 213 | 213 | 213 |
| Laboratory tests | 200 | 200 | 200 |
| Demographics | 117 | 117 | 117 |
| HCPCS procedure codes | 71 | 71 | 71 |
| Numerical tokens* | 10 | 1 | 0 |

\*Discrete tokenization has 10 quantile bin tokens, Continuous Factored has the <NUM> token, and Continuous Fused has no numerical tokens in the vocabulary.

**Table 2:** Per-patient sequence length distribution by tokenization strategy.

| Percentile | Discrete | Cont. Factored | Cont. Fused |
| --- | --- | --- | --- |
| Median | 471 | 471 | 309 |
| 75th | 2,022 | 2,022 | 1,345 |
| 90th | 6,160 | 6,160 | 4,082 |
| 95th | 11,557 | 11,557 | 7,634 |
| 99th | 32,933 | 32,933 | 21,597 |
| Max | 478,666 | 478,666 | 337,214 |

### Next-Token Prediction Performance

Across three independent training runs (**S2 Table**), the discrete model achieved a mean next-token cross-entropy loss of 0.643 ± 0.001, while the continuous fused model reached 0.600 ± 0.002. The Continuous Fused model achieved the final cross-entropy loss of the Discrete model in approximately 30% of the training budget (**Figure 2**A). To examine prediction performance across clinical event types, we stratified loss by the clinical category of each token (**Figure 2**B). The Continuous Fused model outperformed the Discrete baseline across all 11 token categories evaluated. The most pronounced improvements were observed for death (9.1% reduction), ICU events (9.8%), time (8.9%), and medication tokens (7.9%) (**S3 Table**). We evaluated the accuracy of numeric value prediction using nRMSE (**Figure 2**C). The Continuous model demonstrated superior numeric prediction performance across all 200 numeric observations, which encompass laboratory values, vital signs, and time intervals, with a median nRMSE improvement of 30.25% (IQR 15.33%–39.22%). Full results for each type of numeric observation are in **S4 Table**.

**Figure 2:**
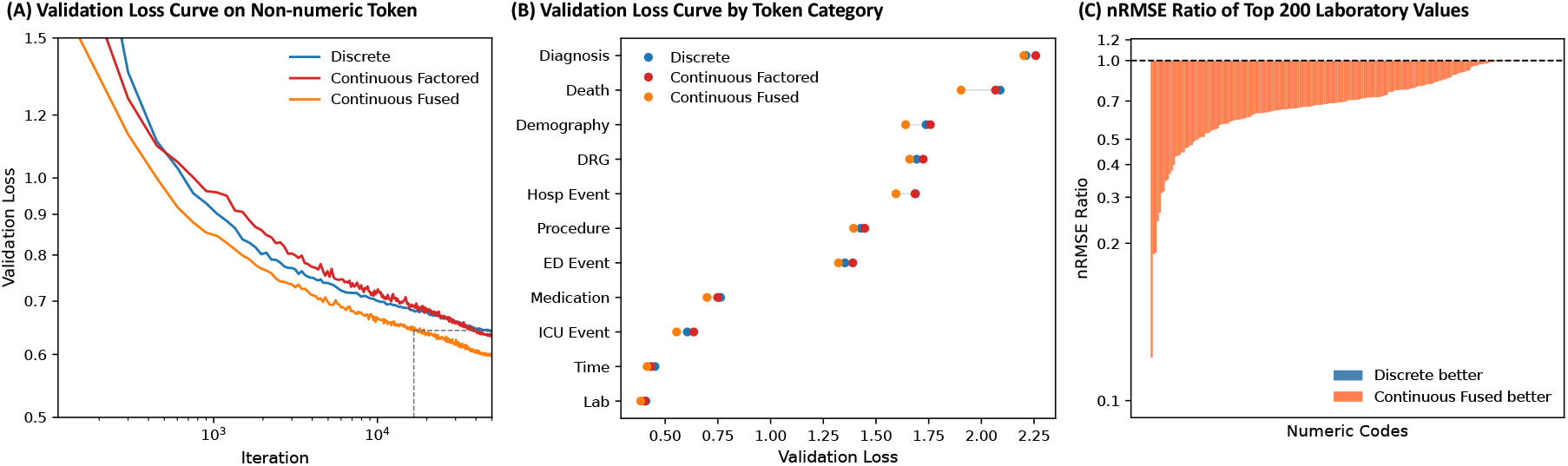
Comparison of performance for non-numeric next-token prediction and next-value prediction. (A) Masked validation loss curves for non-numeric token predictions during training. The model trained with continuous fused tokenization achieved the lowest loss, reaching the final loss of the model trained with discrete tokenization at around 30% of the training iterations (gray line). (B) Validation loss for non-numeric next token prediction by token category. The continuous fused model outperforms the discrete model across all main token categories. (C) Ratio of next-value prediction accuracy between continuous fused and discrete models measured by normalized nRMSE. A value of 1 denotes equivalent nRMSE.

### ED Disposition Prediction

Class prevalences in the evaluation cohort were: Home 57.8%, Admit Hospital 30.8%, Admit ICU 5.4%, Home Unintended 3.6%, Transfer 1.5%, Other 1.0%, and Expired <0.1%. The percentages of simulations that reached a valid disposition were 98.6% for Discrete, 97.2% for Continuous Factored, and 95.2% for Continuous Fused. All three models achieved equivalent Top-1 accuracy of approximately 0.900 for predicting ED disposition at the time of triage across 2,000 encounters (**Figure 3**, Panel A): Continuous Fused 0.901 (95% confidence interval [CI] 0.888 to 0.914), Discrete 0.900 (95% CI 0.887 to 0.913), and Continuous Factored 0.900 (95% CI 0.887 to 0.913). All pairwise comparisons of clinical task performance were evaluated using paired bootstrap tests with Continuous Fused as the reference model. Top-1 accuracy differences were not statistically significant for either Discrete (p = 0.70) or Continuous Factored (p = 0.72).

**Figure 3:**
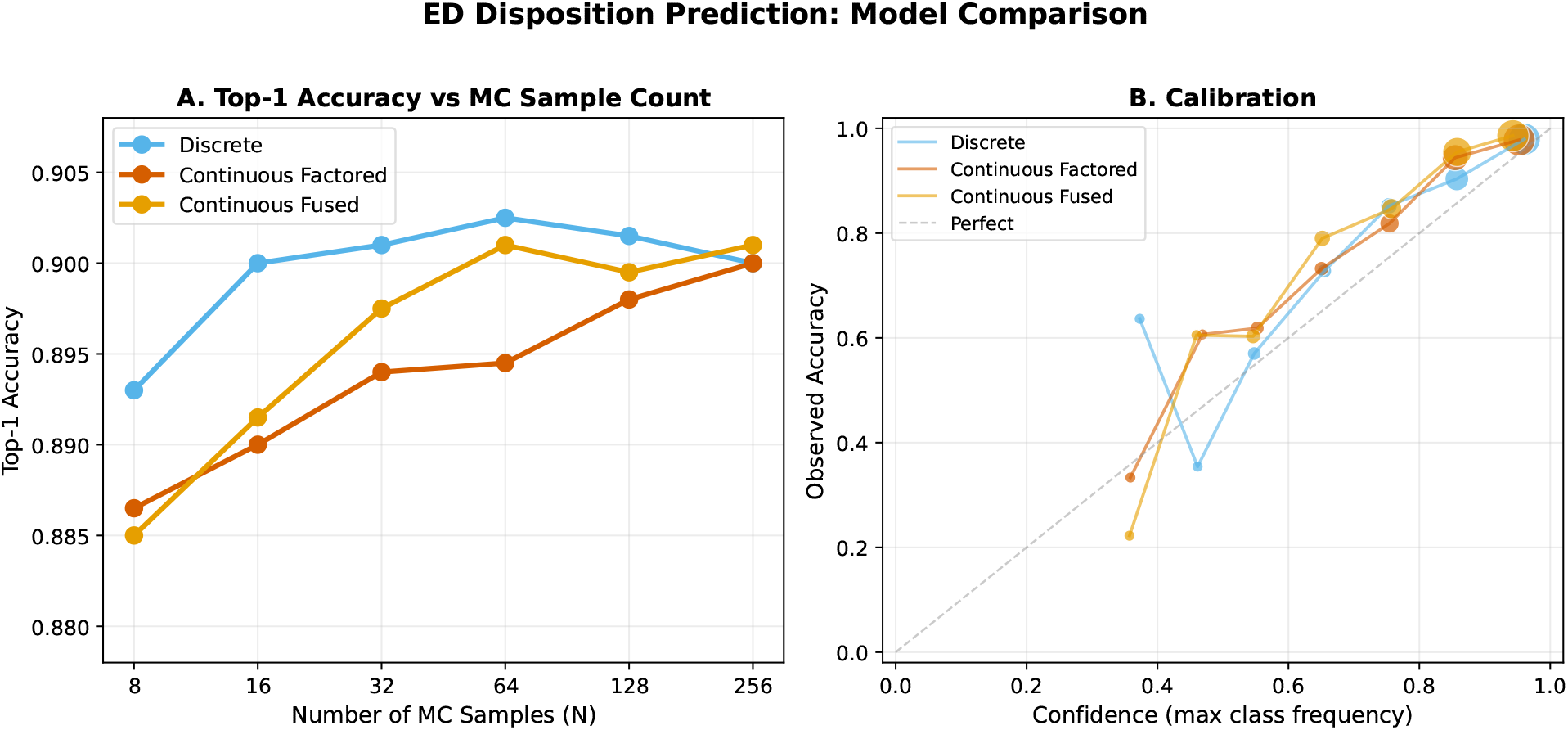
Model performance for ED Disposition prediction at time of triage. (A) Top-1 accuracy by Monte Carlo sample count for all three tokenization variants. All models converge to approximately 0.900 by N = 64. (B) Calibration curves (predicted probability vs. observed frequency) at N = 256 for each model. The Discrete model maintains the best calibration across all sample counts (ECE 0.038 vs. 0.076 for Continuous Fused).

Calibration differed meaningfully across models (**Figure 3**, Panel B). The Discrete model achieved significantly lower ECE (0.038, 95% CI 0.028 to 0.049) than Continuous Fused (0.076, 95% CI 0.064 to 0.088; p < 0.001), with Continuous Factored intermediate (0.053, 95% CI 0.042 to 0.065; p < 0.001 vs. Continuous Fused).

### ICD Code Prediction

From the 2,000 hospital discharge cohort, the numbers of evaluable encounters per task based on ground-truth label availability were 1,999 for ICD, 1,646 for DRG-HCFA, and 1,525 for DRG-APR.

**Figure 4**A summarizes multi-label ICD code prediction performance across 1,999 encounters (97% of simulations produced at least one diagnosis code). The Continuous Fused model achieved significantly higher AU-PRC (0.457, 95% CI 0.448 to 0.465) than both Discrete (0.446, 95% CI 0.438 to 0.455; p < 0.001) and Continuous Factored (0.432, 95% CI 0.424 to 0.440; p < 0.001) in paired bootstrap comparisons (**Table 3**). AU-PRC improved with increasing MC sample count for all models. When stratified by code prevalence (**Table 4**), all models performed best on common codes (AU-PRC: 0.514, 95% CI 0.505 to 0.523 through 0.538, 95% CI 0.529 to 0.547), with performance decreasing for moderate (0.208, 95% CI 0.195 to 0.222 through 0.241, 95% CI 0.227 to 0.255) and rare codes (0.098, 95% CI 0.080 to 0.118 through 0.114, 95% CI 0.095 to 0.135). The Continuous Fused model achieved the highest AU-PRC for common and moderate codes; for rare codes, Discrete and Continuous Fused performed comparably (0.114 vs. 0.114, overlapping CIs).

**Figure 4:**
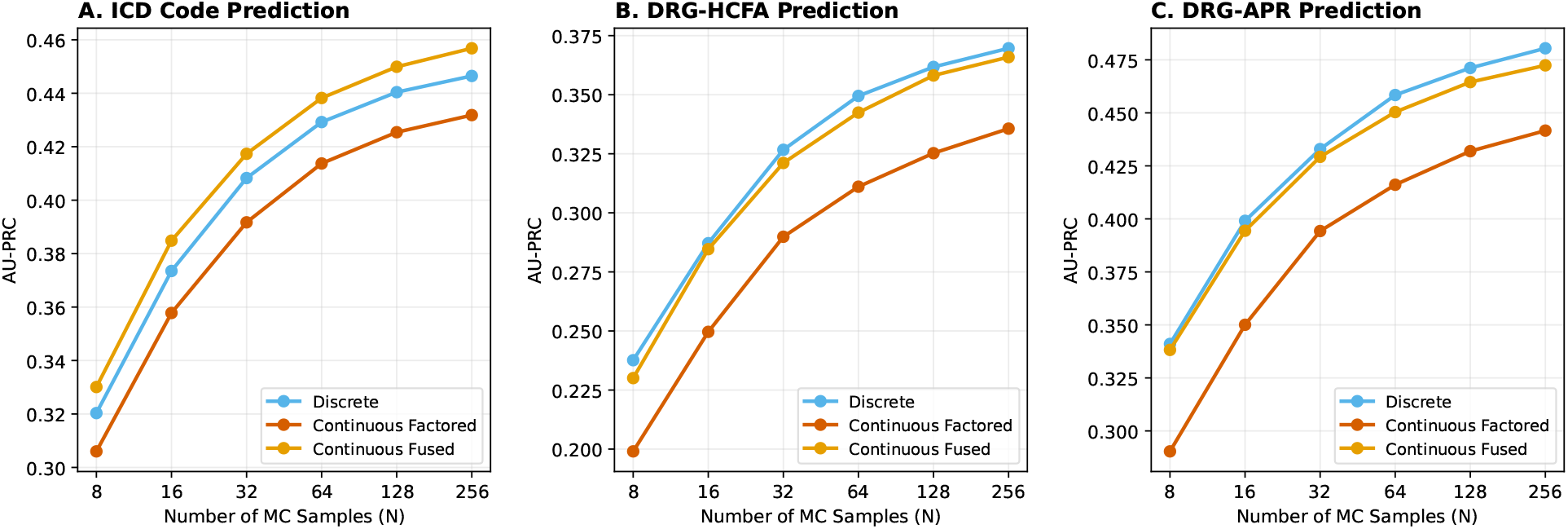
ICD and DRG code prediction performance by tokenization variant. AU-PRC as a function of Monte Carlo sample count for (A) ICD code prediction across 1,999 encounters, (B) DRG-HCFA code prediction across 1,646 encounters, and (C) DRG-APR code prediction across 1,525 encounters. Continuous Fused leads for ICD; Discrete and Continuous Fused converge for both DRG systems. All models improve with increasing sample count.

**Table 3:** Summary of AU-PRC performance across tokenization models for ICD and DRG prediction tasks.

| Task | Model | AU-PRC [95% CI]* | p** |
| --- | --- | --- | --- |
| ICD | Discrete | 0.4465 [0.4380, 0.4551] | <0.001 |
|  | Continuous Factored | 0.4319 [0.4235, 0.4403] | <0.001 |
|  | Continuous Fused | **0.4568 [0.4484, 0.4653]** | ref |
| DRG-HCFA | Discrete | **0.3695 [0.3438, 0.3948]** | 0.49 |
|  | Continuous Factored | 0.3355 [0.3104, 0.3604] | <0.001 |
|  | Continuous Fused | 0.3658 [0.3402, 0.3914] | ref |
| DRG-APR | Discrete | **0.4802 [0.4535, 0.5077]** | 0.19 |
|  | Continuous Factored | 0.4414 [0.4141, 0.4686] | <0.001 |
|  | Continuous Fused | 0.4722 [0.4451, 0.4994] | ref |

\*AU-PRC computed via threshold sweep on Monte Carlo (MC) code frequencies (N=256 samples). 95% bootstrap confidence intervals computed with 10,000 iterations for all tasks. DRG results reported separately for HCFA and APR classification systems. Bold indicates best performance per task. \*\*P-values computed via two-sided paired bootstrap test against Continuous Fused (reference), with significance threshold = 0.05.

### DRG Code Prediction

**Figure 4**B and **Figure 4**C summarize single-label DRG prediction performance for both the HCFA (n = 1,646 encounters) and APR (n = 1,525 encounters) classification systems. DRG completion rates were lower than ICD (60–64% for HCFA, 57–59% for APR). For HCFA, Discrete (0.370, 95% CI 0.344 to 0.395) and Continuous Fused (0.366, 95% CI 0.340 to 0.391) achieved statistically equivalent AU-PRC (p = 0.49), while Continuous Factored (0.336, 95% CI 0.310 to 0.360) was significantly lower (p < 0.001 vs. Continuous Fused). For APR, Discrete (0.480, 95% CI 0.454 to 0.508) and Continuous Fused (0.472, 95% CI 0.445 to 0.499) were again statistically equivalent (p = 0.19), with Continuous Factored (0.441, 95% CI 0.414 to 0.469) significantly lower (p < 0.001 vs. Continuous Fused) (**Table 3**). AU-PRC improved with MC sample count. When stratified by code prevalence (**Table 4**), all models showed higher AU-PRC for common codes relative to rare codes for both classification systems. APR codes were predicted with higher AU-PRC than HCFA codes across all prevalence bins.

**Table 4:**
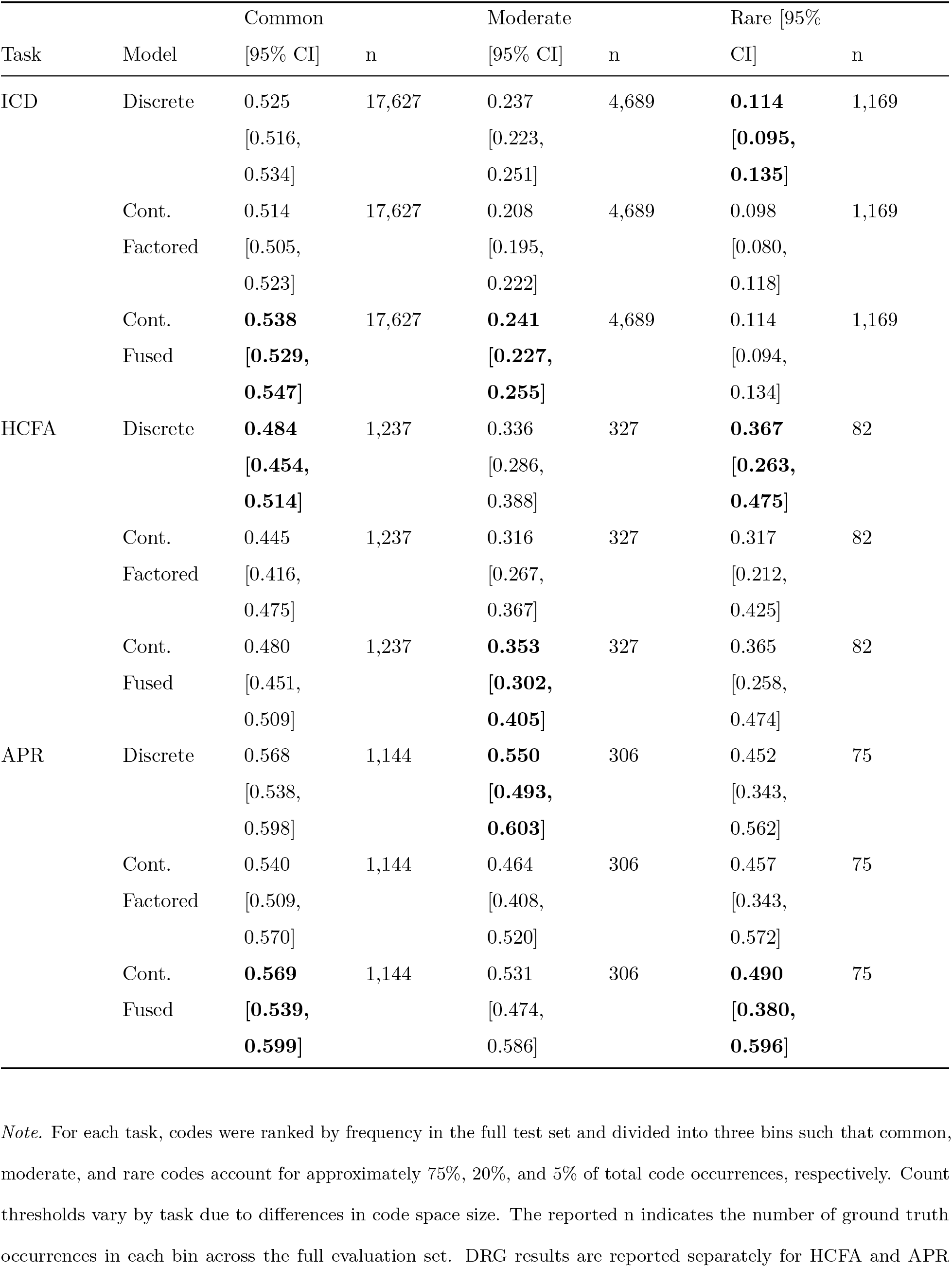
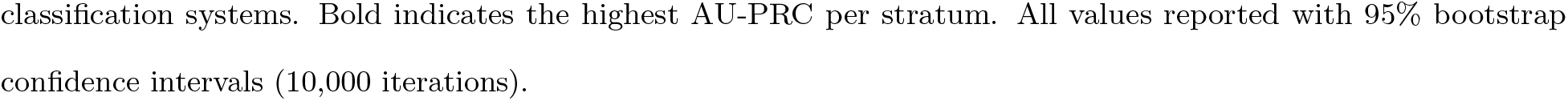
AU-PRC stratified by code prevalence for ICD and DRG prediction tasks.

### Performance versus Monte Carlo Sample Count

Across all tasks and metrics, increasing the number of Monte Carlo simulations improved measured performance. This phenomenon has been observed in prior work [3] but not explained.

This effect arises because many evaluation metrics apply nonlinear transformations to probability estimates (e.g., argmax, logarithm, rank ordering), producing a finite-sample bias that decreases predictably as sample count increases. The mathematical basis for this bias is derived in **S1 Appendix** (see also **S1 Fig**).

This predictable relationship between sample count and performance acts as a scaling law: by fitting the observed trajectory from a small initial sweep, researchers can forecast performance at higher sample counts without running the full experiment (**Figure 5**). Across all 15 metric– variant combinations (5 metrics × 3 tokenization variants), the scaling law predicted 128-sample performance from early sweeps with a median absolute error of 0.22 percentage points (maximum 0.43 pp; **Figure 5**, middle column). Across all three prediction horizons, the median absolute error was 0.16 percentage points (maximum 1.00 pp).

**Figure 5:**
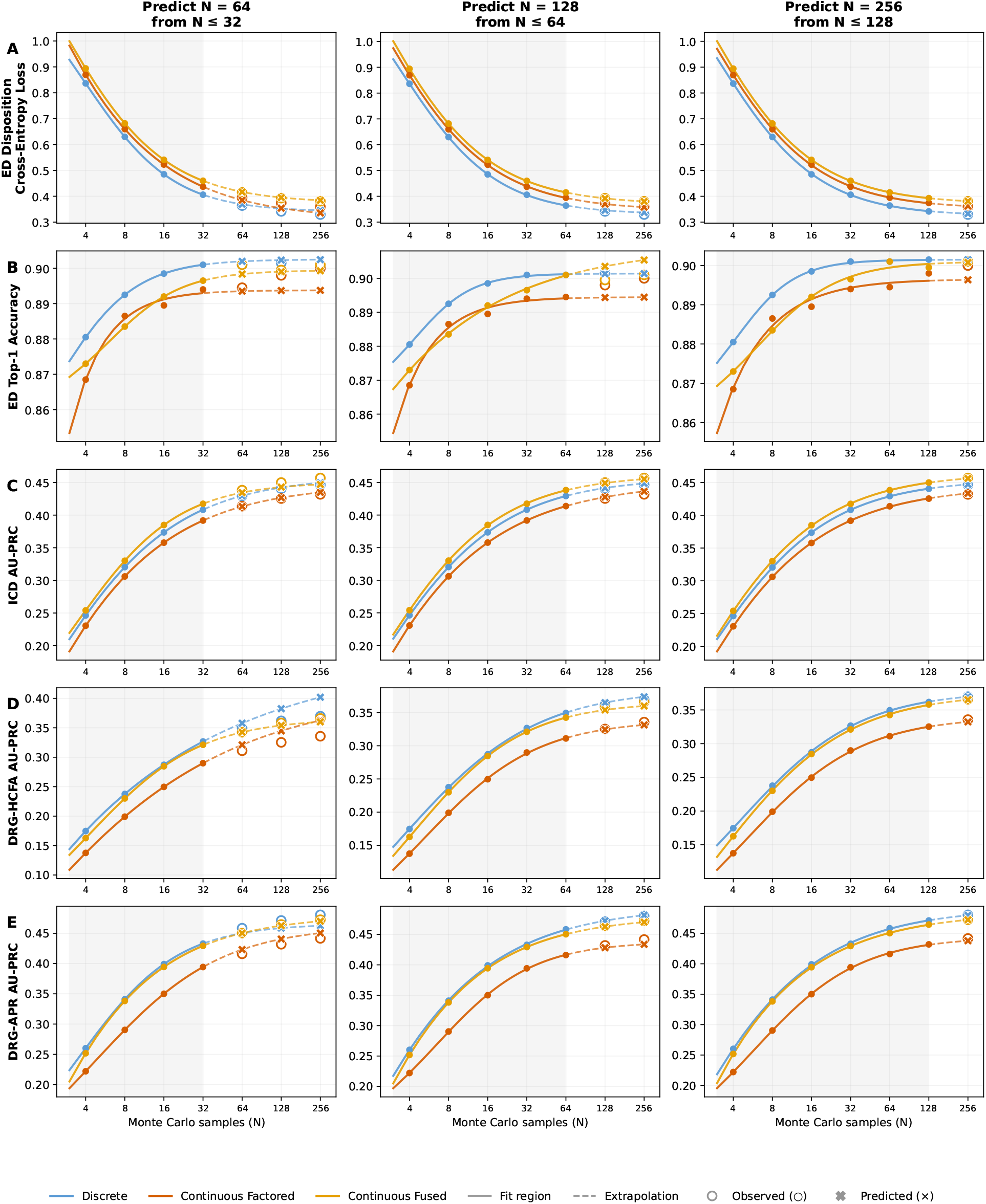
Predicted versus observed evaluation metrics across Monte Carlo sample counts and prediction horizons. Rows show ED cross-entropy loss, ED top-1 accuracy, ICD AU-PRC, DRG-HCFA AU-PRC, and DRG-APR AU-PRC; columns show prediction of N = 64 from N ≤ 32, N = 128 from N ≤ 64, and N = 256 from N ≤ 128. Gray shading indicates the fit region; white indicates extrapolation. Solid lines show fitted scaling-law trajectories in the fit region; dashed lines show extrapolation. Filled circles are observed fit-region values, open circles are observed held-out values, and × markers are scaling-law predictions. Across all 45 task–variant–horizon combinations, median prediction error was 0.16 percentage

## Discussion

We compared three numerical tokenization strategies for a medical event foundation model trained on the MIMIC-IV dataset. The primary finding is that continuous-value tokenization produces substantial gains in training efficiency, numeric prediction accuracy, and sequence compression. The Continuous Fused model converged to the Discrete model’s final cross-entropy loss using only 30% of the training iterations and demonstrated a 30.25% median improvement in nRMSE for next numeric value prediction. Continuous Fused tokenization also reduced median sequence length by 34%, allowing the model to access a larger portion of each patient’s history within a fixed context window.

Clinical task performance varied by task rather than favoring one tokenization strategy uniformly. ICD code prediction was a clear win for Continuous Fused (AU-PRC 0.457 vs. 0.446, p < 0.001), with the advantage present for common and moderate prevalence codes; for rare codes, Discrete and Continuous Fused performed comparably. DRG prediction was statistically equivalent between Continuous Fused and Discrete for both the HCFA (0.366 vs. 0.370, p = 0.49) and APR (0.472 vs. 0.480, p = 0.19) systems. ED disposition top-1 accuracy was equivalent across all models (∼0.900), though the Discrete model achieved better calibration (ECE 0.038 vs. 0.076, p < 0.001).

Current medical event foundation models handle numeric clinical quantities using either categorical value bins, as in ETHOS and Curiosity [3,4], or broader finite categorical event vocabularies, as in Delphi, MOTOR, and CLMBR [5–7]. Our results provide large-scale experimental evidence that continuous-value tokenization offers substantial improvements in training efficiency and numeric prediction accuracy while achieving comparable clinical task performance overall. Combined with reduced sequence length and retained numeric precision, continuous-value tokenization offers practical advantages for medical event foundation models. The approach requires no modifications to standard autoregressive transformer architectures. Lee et al. [22] benchmark numeric representation choices across 28 matched transformers on MIMIC-IV, confirming that code-value fusion produces the most consistent downstream gains. Their one continuous encoder (xVal) underperformed all discrete approaches, which the authors attribute to near-median suppression in the shared multiplicative embedding. Our continuous encoding avoids this failure mode by attaching continuous scalars to observation-specific embeddings rather than scaling a shared placeholder. The evaluation strategies also differ: Lee et al. fit linear probes on frozen representations, while we generate complete clinical trajectories through Monte Carlo sampling.

The Continuous Factored model, which preserves continuous values but maintains the same sequence length as the Discrete baseline, achieved lower validation cross-entropy loss but did not consistently outperform the Discrete model on downstream clinical tasks. This dissociation between pre-training loss and downstream performance is well-documented in the language model literature. Liu et al. demonstrated that pre-training loss cannot fully explain downstream performance and that models with the same pre-training loss can differ meaningfully on downstream tasks [23]. More broadly, recent work has highlighted both the promise and limitations of scaling-law predictions for downstream performance, underscoring the importance of comparing both next-token prediction and task performance when comparing models [24,25].

While previous studies have observed that predictive performance depends on Monte Carlo sample count [3], the reasons for this have not been explored. We provide a mathematical derivation of how finite-sample estimation error propagates through nonlinear evaluation metrics (**S1 Appendix**), explaining the observed performance gains. The resulting scaling law allows researchers to predict performance at an increased sample count from a smaller initial sweep, enabling them to calculate expected gains from doubling compute to identify the point of diminishing returns and avoid wasted time and compute. This becomes increasingly valuable as medical event foundation models scale to larger context windows and evaluation cohorts, where Monte Carlo simulation costs grow substantially.

### Limitations

This study has several limitations. First, the data set is from a single health system and has a high degree of ED and ICU observations. The relative performance of different tokenization strategies may differ in predominantly ambulatory settings. We did not assess whether tokenization effects on prediction performance differ across demographic subgroups (e.g., by sex, age, or race/ethnicity). Second, while we assessed performance in predicting values of all 200 numeric observations, we did not assess performance on long time-horizon numerical outcome predictions. Third, we compared the continuous value tokenization to the discretization method used in published work. If purely discrete tokens are required, different discretization strategies such as adaptive binning, or bins aligned with certain clinical thresholds may improve performance. Fourth, our clinical predictions used 256 Monte Carlo samples per encounter across evaluation cohorts of 2,000 (ED disposition), 1,999 (ICD), 1,646 (DRG-HCFA), and 1,525 (DRG-APR) encounters. While bootstrap confidence intervals provide estimates of uncertainty, these cohort sizes may limit statistical power to detect smaller differences between tokenization strategies, particularly for rare outcomes. Fifth, the interaction between tokenization strategy and model size was not explored; it is possible that larger models or longer training may differentially benefit from continuous-value representations. Sixth, all models used a 512-token context window, compared to the 2,048 tokens in prior works [4], and it remains untested whether tokenization effects differ at longer context windows.

### Future Work

Future work should focus on exploration of how the observed performance differences scale to much larger model sizes, larger training data sets, and longer context windows to understand whether performance differences expand or decrease. Additionally, this tokenization method should be compared in data sets with a larger proportion of ambulatory data and with the addition of other clinical data modalities such as notes, imaging, and waveform data to see how performance changes. Finally, evaluation of training or fine-tuning on smaller data sets should be performed to see if this method could translate to better performance in data-limited settings. Future studies should also evaluate fairness, including whether tokenization strategy interacts with demographic subgroup to affect prediction accuracy.

## Conclusion

We introduce a continuous-value tokenization method that integrates directly into standard transformer architectures and the MEDS data ecosystem, requiring no modifications to the underlying model. Applied to a medical event foundation model trained on MIMIC-IV, this approach reduced median sequence lengths by 34% and training iterations by 70%, and improved numeric prediction accuracy by 30.25%, while achieving comparable clinical task performance. We additionally explain why predictive performance improves with Monte Carlo sample count and derive a scaling law that predicts performance gains from increasing simulation budget. By preserving numerical precision within a more compact representation, continuous-value tokenization offers a practical, computationally efficient path toward more capable medical foundation models.

## Supporting information

TRIPOD AI Checklist

## AI/LLM Disclosure

AI tools (Claude Opus 3.6 and 3.7, Anthropic) were used to assist with manuscript preparation, including drafting, editing, and analysis code development. All AI-generated content was reviewed, verified, and edited by the authors. No AI tools were used for experiment conception, experimental design decisions, or data collection. The authors take full responsibility for the content of the manuscript.

## Data Availability

MIMIC-IV is available through PhysioNet (https://physionet.org/content/mimiciv/) under a credentialed-access Data Use Agreement. Researchers must complete human-subjects research training and agree to the data use restrictions.

## Code Availability

Tokenization code will be made available at (https://github.com/kmacman/Continuous_Value_Tokenization) upon publication.

## Author Contributions

Author contributions are reported using the CRediT (Contributor Roles Taxonomy) framework:

- **Conceptualization:** KAM, IS, AJL
- **Data curation:** KAM, IS
- **Formal analysis:** KAM, IS
- **Investigation:** KAM, IS, HL
- **Methodology:** KAM, AJL, IS
- **Software:** KAM, IS, HL, AJL
- **Supervision:** AJL, ERM, MSI
- **Validation:** KAM, IS, HL
- **Visualization:** KAM, IS
- **Writing – original draft:** KAM, IS
- **Writing – review & editing:** KAM, IS, HL, MSI, ERM, MI, AJL

## Supporting Information

**S1 Checklist. TRIPOD+AI checklist.** Completed TRIPOD+AI reporting checklist with page references for each item.

**S1 Appendix. Monte Carlo estimation bias in generative models.** Mathematical derivation of how finite-sample estimation error propagates through nonlinear evaluation metrics, including metric curvature analysis, scaling law for extrapolating performance, and empirical validation.

**S1 Fig. Effect of metric transformation on Monte Carlo estimation bias.** Computed from 2,000 synthetic encounters (Dirichlet-distributed, 5 classes, 256 MC samples). Panels show (A) cross-entropy loss (convex, overestimates at low N), (B) sampling accuracy (linear, unbiased), (C) top-1 accuracy (concave, underestimates at low N), and (D) aggregate uncertainty (concave, underestimates at low N). Shaded bands show 95% bootstrap confidence intervals.

**S1 Table. Detailed token statistics (Discrete).** Count of unique tokens and total token occurrences by code group for the Discrete tokenization strategy, stratified by training and test splits.

**S2 Table. Per-run validation loss across models.** Final masked validation cross-entropy loss for each of three independent training runs per tokenization strategy.

**S3 Table. Mean group loss by tokenization strategy.** Per-category next-token cross-entropy loss for all 11 clinical event categories.

**S4 Table. RMSE ratio of top 200 numeric observations relative to the Discrete baseline.** Normalized RMSE for each numeric observation across tokenization strategies.

