## Supplementary Materials for "Continuous Value Tokenization Improves Medical Event Foundation Models"

### Supporting Information

Kent A McCann, MD[1,*], Ikgyu Shin, MHS[2,*], Huan Li, PhD[1,2], Davis White[3], Edward R Melnick, MD, MHS[1], Mark Iscoe, MD, MHS[1,2], Andrew J Loza, MD, PhD[2,4]

[1] Department of Emergency Medicine, Yale School of Medicine, New Haven, CT

[2] Department of Biomedical Informatics and Data Science, Yale School of Medicine, New Haven, CT

[3] Epic Systems

[4] Yale New Haven Health System, New Haven, CT

[*] These authors contributed equally to this work.

### S1 Appendix: Monte Carlo Estimation Bias in Generative Models

#### 1 Monte Carlo Estimation in Generative Models

Let a generative model $P_\theta$ be one that learns the joint distribution

$$P_\theta(X_{1:T}) = P(X_{1:T}), \tag{1}$$

where $X$ is a sequence (such as medical events) indexed from 1 to $T$. For any partial history $X_{1:t}$, we can compute the conditional distribution of the future sequence:

$$P_\theta(X_{t+1:T} \mid X_{1:t}). \tag{2}$$

Concatenating the known history $X_{1:t}$ with a sampled future $X_{t+1:T}$ results in a complete sample. If Eq. (1) holds, Eq. (2) represents a valid factorization of the full distribution.

Given a binary outcome $Y$ computable from the future sequence $X_{t+1:T}$, we marginalize over all possible futures to obtain the true conditional probability $p$:

$$p = P_\theta(Y \mid X_{1:t}) = \sum_{X_{t+1:T}} \mathbf{1}\{Y \in X_{t+1:T}\}\, P_\theta(X_{t+1:T} \mid X_{1:t}). \tag{3}$$

Marginalizing over the true set of all future sequences is often intractable, so we approximate $p$ using a Monte Carlo estimator $\widehat{p}_N$ where $N$ is the number of sequences used in the estimate. The estimator is defined as:

$$\widehat{p}_N = \frac{1}{N}\sum_{i=1}^{N} \mathbf{1}\{Y \in X_{t+1:T}^{(i)}\}. \tag{4}$$

This estimator $\widehat{p}_N$ is the central object of our analysis regarding bias under finite sampling.

### 2 Source of Bias Under Finite Sample Size

We now analyze the behavior of $\widehat{p}_N$ as defined in Eq. (4). We assume the estimator is consistent ($\lim_{N\to\infty} \widehat{p}_N = p$) and unbiased with respect to the raw probability, such that the estimation error $\epsilon(N) = \widehat{p}_N - p$ satisfies:

$$\mathbb{E}[\epsilon(N)] = 0, \quad \text{and} \quad \mathbb{E}[\epsilon(N)^2] = \mathrm{Var}(\widehat{p}_N) = \frac{\sigma^2}{N}, \tag{5}$$

where $\sigma^2$ is the per-sample variance (e.g., $p(1-p)$ for Bernoulli trials).

The central question is: under what conditions is a *transformation* of the estimator, $f(\widehat{p}_N)$, an unbiased estimate of the transformed parameter $f(p)$?

$$\mathbb{E}[f(\widehat{p}_N)] \stackrel{?}{=} f(p) \tag{6}$$

Transformations of the estimator are used for all downstream metrics. We analyze this using the Delta Method, approximating $f(\widehat{p}_N)$ with a second-order Taylor expansion around the true parameter $p$. Substituting $\widehat{p}_N = p + \epsilon$:

$$f(p+\epsilon) \approx f(p) + f'(p)\epsilon + \frac{1}{2} f''(p)\epsilon^2. \tag{7}$$

Taking the expectation $\mathbb{E}[\cdot]$ with respect to the sampling distribution:

$$\mathbb{E}[f(\widehat{p}_N)] \approx f(p) + f'(p)\underbrace{\mathbb{E}[\epsilon]}_{0} + \frac{1}{2} f''(p)\underbrace{\mathbb{E}[\epsilon^2]}_{\sigma^2/N}. \tag{8}$$

This yields the **Estimator Bias Equation**:

$$\boxed{\mathbb{E}[f(\hat{p}_N)] \approx f(p) + \frac{\sigma^2}{2N} f''(p)} \tag{9}$$

This relation highlights the conditions required for the estimator to be unbiased ($\mathbb{E}[f(\hat{p}_N)] = f(p)$):

- **Linearity:** $f''(p) = 0$.
- **Zero Variance:** $\sigma^2 = 0$ (deterministic outcomes).
- **Infinite Samples:** As $N \to \infty$, the bias term vanishes.

#### Impact of Metric Curvature

The direction of the bias is determined entirely by the curvature $f''(p)$. If $f$ is convex ($f''(p) > 0$), the estimator will systematically overestimate the true value. An example is cross entropy loss because $-\log(p)$ is a convex function. We empirically see that $\mathbb{E}[f(\hat{p}_N)]$ decreases with increasing $N$. By choosing a related metric that is linear, we can demonstrate how an estimator can be unbiased. Instead of computing $f = -\log(\hat{p}_{N,i})$, we instead compute $f = \text{avg}(\hat{p}_{N,i})$ (collecting the probability assigned to the true class, not the negative log of the probability), we find that $\mathbb{E}[f(\hat{p}_N)]$ is unbiased even at small $N$. Top-K Accuracy provides an example of a concave characteristic although it is not smoothly differentiable. Consider the impact of symmetric noise $\pm|\epsilon|$. Let $\Delta_+ = f(p + |\epsilon|) - f(p)$ and $\Delta_- = f(p - |\epsilon|) - f(p)$. If $\Delta_+ + \Delta_- < 0$, the function is concave. This is seen in functions like Top-K Accuracy under the condition when the correct value is selected most of the time. Positive noise for the correct class probability is unlikely to change the output of $f$ (because it was already selected, $|f(p + |\epsilon|) - f(p)| \approx 0$) whereas negative noise could cause the estimate to switch to an incorrect class. Similar behavior is seen in Precision Recall Area Under the Curve and Receiver Operator Characteristic Area Under the Curve. It is more likely that noise will break correct ordering than improve the ordering. If the magnitudes of these quantities are equal, the estimator will not be biased. These behaviors are illustrated empirically in Figure S1.

#### Estimating Performance at Larger Monte Carlo Sample Numbers

We can use the dependence of bias on $N$ from Eq. (9) to extrapolate expected metric performance and estimate the expected improvement with additional Monte Carlo samples.

Given a candidate sample size $N^*$, we first perform bootstrap resampling on the existing completions to estimate the metric $\hat{f}(N)$ at a series of intermediate values $N_1, N_2, \cdots, N^*$. This yields an empirical curve of metric performance as a function of sample count.

While the Estimator Bias Equation predicts bias decaying as $1/N$, this first-order approximation does not account for higher-order derivatives of $f$, or for the fact that metrics are bounded (i.e. ROC AUC cannot be less than zero and chance performance is 0.5). We therefore fit a curve of the form:

$$\hat{f}(N) = f_\infty - \frac{c}{a + N^b}, \tag{10}$$

where $f_\infty$ is the estimated asymptotic performance, $c = a(f_\infty - f_{\text{floor}})$ encodes the gap between the asymptote and the chance-level minimum performance $f_{\text{floor}}$, $a > 0$ is a constant, and $b > 0$ controls the steepness of convergence. We fit these parameters by nonlinear least squares to the bootstrap curve and extrapolate to larger values of $N$ than $N^*$.

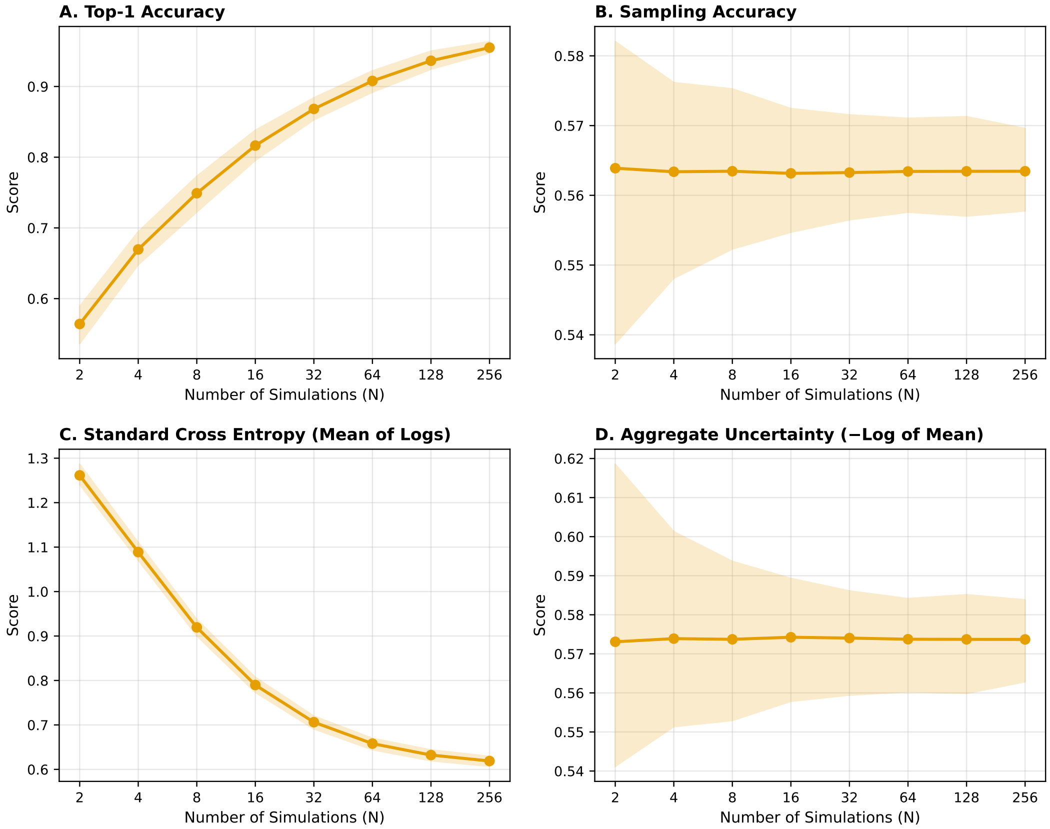


**Figure S1:** Effect of metric transformation on Monte Carlo estimation bias, computed from 2,000 synthetic encounters with 256 simulations each. (A) Top-1 accuracy is concave: positive noise rarely changes a correct prediction, but negative noise can flip it, producing systematic underestimation at low $N$ that improves with more samples. (B) Sampling accuracy (mean predicted probability of the true class) is linear in $\hat{p}_N$ and therefore unbiased at all $N$. (C) Standard cross-entropy (mean of $-\log \hat{p}_{N,i}$) is convex: Jensen's inequality guarantees systematic overestimation (worse loss) that decreases as $N$ grows. (D) Aggregate uncertainty ($-\log(\text{mean}(\hat{p}_{N,i}))$) applies the logarithm after averaging, making it approximately linear and nearly unbiased. Shaded regions show 95% confidence bands.

**Table S1:** Detailed Token Statistics (Discrete). Count of unique tokens and total token occurrences by code group, stratified by training and test splits.

| Code Group | #Unique (Train) | Count (Train) | #Unique (Test) | Count (Test) | #Unique (Total) | Count (Total) |
|---|---|---|---|---|---|---|
| LAB | 200 | 186,378,038 | 200 | 23,141,976 | 200 | 209,520,014 |
| ATC | 84 | 53,648,340 | 80 | 6,753,233 | 84 | 60,401,573 |
| ATC_4 | 12 | 53,648,340 | 12 | 6,753,233 | 12 | 60,401,573 |
| ATC_SFX | 216 | 53,648,340 | 188 | 6,753,233 | 216 | 60,401,573 |
| MEDICATION | 68 | 53,648,340 | 62 | 6,753,233 | 68 | 60,401,573 |
| TIME_UNDER_24 | 1 | 50,354,125 | 1 | 6,272,374 | 1 | 56,626,499 |
| Q6 | 1 | 40,753,296 | 1 | 5,053,110 | 1 | 45,806,406 |
| Q5 | 1 | 30,761,612 | 1 | 3,803,557 | 1 | 34,565,169 |
| Q2 | 1 | 30,162,363 | 1 | 3,765,422 | 1 | 33,927,785 |
| Q4 | 1 | 30,110,352 | 1 | 3,735,680 | 1 | 33,846,032 |
| Q7 | 1 | 29,842,322 | 1 | 3,726,884 | 1 | 33,569,206 |
| Q8 | 1 | 28,694,781 | 1 | 3,579,869 | 1 | 32,274,650 |
| Q3 | 1 | 27,602,565 | 1 | 3,436,641 | 1 | 31,039,206 |
| Q9 | 1 | 25,917,914 | 1 | 3,239,167 | 1 | 29,157,081 |
| Q1 | 1 | 8,692,244 | 1 | 1,066,858 | 1 | 9,759,102 |
| VITAL | 7 | 7,494,216 | 7 | 929,423 | 7 | 8,423,639 |
| Q10 | 1 | 5,926,139 | 1 | 745,329 | 1 | 6,671,468 |
| ICD_CM_CHAR_13 | 1,810 | 5,798,202 | 1,619 | 733,851 | 1,819 | 6,532,053 |
| ICD_CM_CHAR_45 | 188 | 5,475,843 | 165 | 693,677 | 188 | 6,169,520 |
| ICD_PCS | 35 | 4,767,994 | 35 | 599,865 | 35 | 5,367,859 |
| TIME_OVER_24 | 1 | 3,286,504 | 1 | 418,706 | 1 | 3,705,210 |
| TRIAGE | 8 | 2,596,139 | 8 | 322,823 | 8 | 2,918,962 |
| BP Systolic | 6 | 2,260,497 | 6 | 289,317 | 6 | 2,549,814 |
| BP Diastolic | 6 | 2,260,497 | 6 | 289,317 | 6 | 2,549,814 |
| TRANSFER_TO | 92 | 1,927,868 | 89 | 242,040 | 92 | 2,169,908 |
| Weight | 1 | 1,648,818 | 1 | 210,174 | 1 | 1,858,992 |
| BMI | 1 | 1,536,926 | 1 | 195,779 | 1 | 1,732,705 |

| Code Group | #Unique (Train) | Count (Train) | #Unique (Test) | Count (Test) | #Unique (Total) | Count (Total) |
|---|---|---|---|---|---|---|
| PROCEDURE | 318 | 1,264,979 | 306 | 155,403 | 318 | 1,420,382 |
| ICD_CM_CHAR_67 | 161 | 1,113,643 | 128 | 141,233 | 161 | 1,254,876 |
| AGE | 92 | 761,762 | 86 | 95,911 | 92 | 857,673 |
| Height | 1 | 647,828 | 1 | 82,628 | 1 | 730,456 |
| DRG | 1,848 | 608,512 | 1,633 | 76,864 | 1,853 | 685,376 |
| HOSPITAL_ADMISSION | 70 | 435,803 | 66 | 54,897 | 72 | 490,700 |
| HOSPITAL_DISCHARGE | 14 | 435,803 | 14 | 54,897 | 14 | 490,700 |
| RACE | 33 | 435,803 | 33 | 54,897 | 33 | 490,700 |
| LANGUAGE | 25 | 435,175 | 25 | 54,825 | 25 | 490,000 |
| INSURANCE | 5 | 428,291 | 5 | 53,964 | 5 | 482,255 |
| MARITAL | 4 | 424,917 | 4 | 53,564 | 4 | 478,481 |
| ED_TRANSPORT | 5 | 339,907 | 5 | 42,189 | 5 | 382,096 |
| ED_DISPOSITION | 8 | 339,907 | 8 | 42,189 | 8 | 382,096 |
| ED_OUT | 1 | 298,783 | 1 | 37,669 | 1 | 336,452 |
| ED_REGISTRATION | 1 | 298,783 | 1 | 37,669 | 1 | 336,452 |
| GENDER | 2 | 291,702 | 2 | 36,463 | 2 | 328,165 |
| MEDS_BIRTH | 1 | 291,702 | 1 | 36,463 | 1 | 328,165 |
| SOS | 1 | 291,702 | 1 | 36,463 | 1 | 328,165 |
| HCPCS | 70 | 138,013 | 38 | 17,417 | 70 | 155,430 |
| ICU_ADMISSION | 16 | 75,734 | 14 | 9,293 | 17 | 85,027 |
| ICU_DISCHARGE | 16 | 75,723 | 14 | 9,292 | 17 | 85,015 |
| MEDS_DEATH | 1 | 30,670 | 1 | 3,831 | 1 | 34,501 |

**Table S2:** Per-run validation loss across models. Final masked validation cross-entropy loss for each of three independent training runs per tokenization strategy.

| Model | Run 1 | Run 2 | Run 3 |
|---|---|---|---|
| Discrete | 0.643 | 0.643 | 0.644 |
| Continuous Fused | 0.602 | 0.598 | 0.599 |
| Continuous Factored | 0.635 | 0.633 | 0.634 |

**Table S3:** Mean group loss by tokenization strategy. Per-category next-token cross-entropy loss for all 11 clinical event categories.

| Group | Discrete | Continuous Fused | Continuous Factored |
|---|---|---|---|
| Lab | 0.41 | 0.38 | 0.40 |
| Time | 0.45 | 0.41 | 0.43 |
| ICU Event | 0.61 | 0.55 | 0.64 |
| Medication | 0.76 | 0.70 | 0.75 |
| ED Event | 1.35 | 1.32 | 1.39 |
| Procedure | 1.43 | 1.39 | 1.45 |
| Hosp Event | 1.69 | 1.59 | 1.69 |
| Demography | 1.74 | 1.64 | 1.76 |
| DRG | 1.69 | 1.66 | 1.72 |
| Death | 2.09 | 1.90 | 2.07 |
| Diagnosis | 2.21 | 2.20 | 2.26 |

**Table S4:** RMSE ratio of top 200 numeric observations relative to the Discrete baseline. Values below 1.0 indicate the continuous model outperforms Discrete. Sorted by Continuous Fused ratio.

| Lab Name | Cont. Fused | Cont. Factored | Prevalence |
|---|---|---|---|
| Post Filter Replacement Rate | 0.0731 | 0.2981 | 0.13% |
| eAG | 0.1817 | 0.2413 | 0.11% |
| MCHC | 0.1839 | 0.2047 | 0.82% |
| ALT | 0.2431 | 0.2424 | 0.07% |
| MCV | 0.2620 | 0.2940 | 1.78% |
| MCHC | 0.3116 | 0.3240 | 0.96% |
| AST | 0.3154 | 0.2589 | 0.07% |
| PTT | 0.3458 | 0.3903 | 0.17% |
| HCO3 (serum) | 0.3504 | 0.4198 | 0.25% |
| pCO2 | 0.3680 | 0.4029 | 0.30% |
| Red Blood Cells | 0.3791 | 0.4879 | 1.78% |
| Absolute Monocyte Count | 0.3983 | 0.4938 | 0.36% |
| RDW-SD | 0.4293 | 0.4908 | 0.96% |
| Neutrophils | 0.4318 | 0.4887 | 0.69% |
| Creatine Kinase (CK) | 0.4356 | 0.4511 | 0.14% |
| Sodium | 0.4455 | 0.4908 | 1.77% |
| pH | 0.4457 | 0.4620 | 0.32% |
| Sodium (serum) | 0.4628 | 0.4932 | 0.27% |
| Cholesterol, LDL, Calculated | 0.4644 | 0.4958 | 0.12% |
| Calculated Total CO2 | 0.4726 | 0.5054 | 0.30% |
| Hemoglobin | 0.4752 | 0.5066 | 1.79% |
| Anion gap | 0.4892 | 0.5279 | 0.25% |
| Phosphorous | 0.4953 | 0.5125 | 0.24% |
| Trans Membrane Pressure | 0.4997 | 0.5227 | 0.11% |
| Creatinine | 0.5078 | 0.5497 | 1.85% |
| GI #1 Tube Mark (CM) | 0.5094 | 0.5277 | 0.07% |
| Potassium (whole blood) | 0.5106 | 0.5150 | 0.07% |
| Lactate | 0.5268 | 0.5197 | 0.29% |
| Pressure Drop | 0.5271 | 0.5175 | 0.11% |
| Potassium (serum) | 0.5319 | 0.5694 | 0.27% |
| Effluent Pressure | 0.5393 | 0.5978 | 0.15% |
| MCH | 0.5405 | 0.6032 | 1.78% |
| Pulmonary Artery Pressure systolic | 0.5440 | 0.5996 | 0.17% |
| Lymphocytes | 0.5455 | 0.5870 | 0.69% |
| Urea Nitrogen | 0.5509 | 0.6264 | 1.80% |
| Bilirubin, Total | 0.5668 | 0.6325 | 0.68% |
| Central Venous Pressure | 0.5701 | 0.5456 | 0.43% |
| Mean Airway Pressure | 0.5719 | 0.6036 | 0.35% |
| Absolute Neutrophil Count | 0.5738 | 0.6251 | 0.38% |
| Calcium non-ionized | 0.5738 | 0.6160 | 0.23% |

| Lab Name | Cont. Fused | Cont. Factored | Prevalence |
|---|---|---|---|
| Bicarbonate | 0.5823 | 0.6247 | 1.69% |
| ART BP Diastolic | 0.5871 | 0.6145 | 0.17% |
| Resistance Exp | 0.5909 | 0.6311 | 0.08% |
| RDW | 0.5927 | 0.6614 | 1.78% |
| WBC | 0.5941 | 0.5775 | 0.23% |
| Non Invasive Blood Pressure mean | 0.5956 | 0.5838 | 2.31% |
| Citrate (ACD-A) | 0.5977 | 0.7395 | 0.13% |
| Ionized Calcium | 0.5995 | 0.6128 | 0.13% |
| Pinsp (Hamilton) | 0.6053 | 0.6543 | 0.08% |
| Chloride | 0.6056 | 0.6450 | 1.74% |
| Phosphate | 0.6086 | 0.6419 | 1.21% |
| RCexp (Measured Time Constant) | 0.6199 | 0.6263 | 0.08% |
| Base Excess | 0.6207 | 0.6505 | 0.30% |
| Monocytes | 0.6228 | 0.6623 | 0.69% |
| Plateau Pressure | 0.6280 | 0.6555 | 0.13% |
| SvO2 | 0.6291 | 0.6321 | 0.10% |
| Heart Rate | 0.6299 | 0.6662 | 3.77% |
| Pulmonary Artery Pressure diastolic | 0.6301 | 0.6944 | 0.17% |
| Resistance Insp | 0.6311 | 0.6522 | 0.08% |
| Lactate Dehydrogenase (LD) | 0.6335 | 0.6820 | 0.28% |
| Platelet Count | 0.6339 | 0.6845 | 1.80% |
| PTT | 0.6383 | 0.6403 | 0.70% |
| Cholesterol, HDL | 0.6395 | 0.6841 | 0.14% |
| Albumin | 0.6434 | 0.6834 | 0.44% |
| PT | 0.6439 | 0.6515 | 0.76% |
| Hemoglobin | 0.6458 | 0.6274 | 0.26% |
| Hematocrit | 0.6477 | 0.6874 | 1.86% |
| Cardiac Output (CCO) | 0.6507 | 0.6867 | 0.08% |
| L | 0.6511 | 0.6562 | 1.13% |
| EtCO2 | 0.6512 | 0.6612 | 0.07% |
| Calcium, Total | 0.6530 | 0.6974 | 1.28% |
| Arterial Blood Pressure systolic | 0.6548 | 0.6716 | 1.33% |
| Anion Gap | 0.6557 | 0.6783 | 1.69% |
| ART BP Systolic | 0.6562 | 0.6699 | 0.17% |
| Dialysate Rate | 0.6571 | 0.8068 | 0.14% |
| Replacement Rate | 0.6589 | 0.8062 | 0.14% |
| Non Invasive Blood Pressure systolic | 0.6605 | 0.6755 | 2.31% |
| Myelocytes | 0.6646 | 0.6675 | 0.10% |
| White Blood Cells | 0.6651 | 0.7089 | 1.78% |
| Ventilator Tank #1 | 0.6653 | 0.6899 | 0.10% |
| Compliance | 0.6659 | 0.6807 | 0.08% |
| Potassium | 0.6663 | 0.6883 | 1.78% |
| % Hemoglobin A1c | 0.6693 | 0.6739 | 0.13% |
| Access Pressure | 0.6702 | 0.7165 | 0.15% |

| Lab Name | Cont. Fused | Cont. Factored | Prevalence |
|---|---|---|---|
| pO2 | 0.6735 | 0.7006 | 0.30% |
| Inspiratory Time | 0.6756 | 0.7238 | 0.18% |
| Ventilator Tank #2 | 0.6764 | 0.7242 | 0.10% |
| Specific Gravity | 0.6768 | 0.6859 | 0.36% |
| Daily Weight | 0.6781 | 0.6568 | 0.13% |
| Respiratory Rate | 0.6813 | 0.6955 | 3.72% |
| Pminimum | 0.6818 | 0.7021 | 0.08% |
| Alkaline Phosphatase | 0.6857 | 0.7351 | 0.69% |
| Flow Rate (L/min) | 0.6867 | 0.7071 | 0.15% |
| Cuff Pressure | 0.6868 | 0.6732 | 0.07% |
| Glucose finger stick (range 70-100) | 0.6873 | 0.6977 | 0.44% |
| Uric Acid | 0.6897 | 0.7012 | 0.08% |
| Glucose (whole blood) | 0.6911 | 0.5195 | 0.08% |
| Glucose | 0.6923 | 0.6799 | 0.12% |
| Apnea Interval | 0.6937 | 0.7435 | 0.33% |
| Vti High | 0.6957 | 0.7356 | 0.33% |
| Cholesterol, Total | 0.6993 | 0.6268 | 0.14% |
| Free Calcium | 0.7052 | 0.7096 | 0.16% |
| Current Goal | 0.7052 | 0.7005 | 0.14% |
| Oxygen Saturation | 0.7052 | 0.7231 | 0.10% |
| Absolute Basophil Count | 0.7064 | 0.7804 | 0.36% |
| Hourly Patient Fluid Removal | 0.7065 | 0.7441 | 0.15% |
| PEEP set | 0.7083 | 0.7853 | 0.37% |
| Alkaline Phosphate | 0.7092 | 0.6449 | 0.07% |
| Triglycerides | 0.7137 | 0.6714 | 0.14% |
| Eosinophils | 0.7165 | 0.7336 | 0.69% |
| Minute Volume | 0.7178 | 0.7324 | 0.35% |
| Bands | 0.7212 | 0.7148 | 0.11% |
| H | 0.7221 | 0.7252 | 1.14% |
| INR(PT) | 0.7227 | 0.7291 | 0.76% |
| Blood Flow (ml/min) | 0.7239 | 0.7477 | 0.14% |
| Inspired O2 Fraction | 0.7241 | 0.7346 | 0.49% |
| pH | 0.7247 | 0.7214 | 0.36% |
| Potassium, Whole Blood | 0.7248 | 0.7100 | 0.14% |
| Immature Granulocytes | 0.7262 | 0.5734 | 0.29% |
| Magnesium | 0.7310 | 0.7451 | 1.26% |
| Glucose | 0.7346 | 0.7551 | 1.56% |
| Return Pressure | 0.7388 | 0.7608 | 0.15% |
| Creatinine, Urine | 0.7576 | 0.7484 | 0.10% |
| Minute Volume Alarm - Low | 0.7586 | 0.8115 | 0.33% |
| C-Reactive Protein | 0.7601 | 0.7598 | 0.08% |
| Thyroid Stimulating Hormone | 0.7707 | 0.7784 | 0.17% |
| PSV Level | 0.7707 | 0.7650 | 0.18% |
| O2 Flow | 0.7712 | 0.7938 | 0.33% |

| Lab Name | Cont. Fused | Cont. Factored | Prevalence |
|---|---|---|---|
| Tidal Volume (set) | 0.7731 | 0.7979 | 0.19% |
| Total PEEP Level | 0.7732 | 0.8037 | 0.11% |
| O2 saturation pulseoxymetry | 0.7739 | 0.7836 | 3.69% |
| Metamyelocytes | 0.7740 | 0.7655 | 0.10% |
| Basophils | 0.7751 | 0.7853 | 0.69% |
| Globulin | 0.7823 | 0.7451 | 0.07% |
| Arterial Blood Pressure Alarm - Low | 0.7844 | 0.7914 | 0.12% |
| Cerebral Perfusion Pressure | 0.7874 | 0.8142 | 0.07% |
| Paw High | 0.7946 | 0.8327 | 0.33% |
| Peak Insp. Pressure | 0.7989 | 0.8231 | 0.34% |
| Minute Volume Alarm - High | 0.7991 | 0.8383 | 0.33% |
| Fspn High | 0.8032 | 0.8291 | 0.31% |
| Cholesterol Ratio (Total/HDL) | 0.8055 | 0.8073 | 0.14% |
| Protein, Total | 0.8122 | 0.7021 | 0.09% |
| Blood Temperature CCO (C) | 0.8130 | 0.8411 | 0.07% |
| Atypical Lymphocytes | 0.8141 | 0.8005 | 0.09% |
| Resp Alarm - High | 0.8178 | 0.8317 | 0.35% |
| Heparin Dose (per hour) | 0.8265 | 0.8296 | 0.08% |
| Filter Pressure | 0.8288 | 0.8493 | 0.15% |
| I | 0.8366 | 0.8865 | 1.04% |
| Expiratory Ratio | 0.8375 | 0.8538 | 0.10% |
| Alanine Aminotransferase (ALT) | 0.8465 | 0.8599 | 0.78% |
| Asparate Aminotransferase (AST) | 0.8473 | 0.8787 | 0.77% |
| Respiratory Rate (spontaneous) | 0.8546 | 0.8409 | 0.34% |
| Epithelial Cells | 0.8586 | 0.8429 | 0.19% |
| Intra Cranial Pressure | 0.8643 | 0.8547 | 0.11% |
| Creatine Kinase, MB Isoenzyme | 0.8794 | 0.8921 | 0.09% |
| Lipase | 0.8829 | 0.8843 | 0.13% |
| RBC | 0.8872 | 0.8810 | 0.20% |
| Arterial Blood Pressure mean | 0.8974 | 0.8596 | 1.33% |
| Non-Invasive Blood Pressure Alarm - High | 0.8987 | 0.8989 | 0.27% |
| Magnesium | 0.9049 | 0.8679 | 0.25% |
| WBC | 0.9064 | 0.8998 | 0.19% |
| ART BP Mean | 0.9148 | 0.8821 | 0.17% |
| Absolute Lymphocyte Count | 0.9164 | 0.9213 | 0.36% |
| PBP (Prefilter) Replacement Rate | 0.9424 | 0.9432 | 0.13% |
| Respiratory Rate (Set) | 0.9504 | 0.9624 | 0.20% |
| Heart rate Alarm - High | 0.9547 | 0.9609 | 0.36% |
| Pulmonary Artery Pressure mean | 0.9557 | 0.9679 | 0.17% |
| Temperature Celsius | 0.9646 | 0.9697 | 0.17% |
| Tidal Volume (spontaneous) | 0.9688 | 0.9699 | 0.18% |
| Ultrafiltrate Output | 0.9701 | 0.8686 | 0.16% |
| Ferritin | 0.9738 | 0.9698 | 0.08% |
| Absolute Eosinophil Count | 0.9744 | 1.0175 | 0.36% |

| Lab Name | Cont. Fused | Cont. Factored | Prevalence |
|---|---|---|---|
| Non Invasive Blood Pressure diastolic | 0.9846 | 0.9876 | 2.31% |
| Temperature Fahrenheit | 0.9879 | 0.9927 | 0.88% |
| Inspiratory Ratio | 0.9898 | 1.0000 | 0.10% |
| Respiratory Rate (Total) | 0.9911 | 0.9980 | 0.33% |
| SpO2 Desat Limit | 0.9929 | 0.9958 | 0.35% |
| Tidal Volume (observed) | 0.9935 | 0.9998 | 0.35% |
| O2 Saturation Pulseoxymetry Alarm - Low | 0.9936 | 0.9972 | 0.36% |
| Arterial Blood Pressure diastolic | 0.9945 | 0.9998 | 1.33% |
| Inspired Gas Temp. | 0.9952 | 0.9999 | 0.24% |
| Arterial CO2 Pressure | 0.9960 | 0.9999 | 0.18% |
| Arterial O2 pressure | 0.9962 | 1.0000 | 0.18% |
| Arterial Base Excess | 0.9962 | 0.9999 | 0.18% |
| PH (Arterial) | 0.9963 | 1.0000 | 0.19% |
| TCO2 (calc) Arterial | 0.9963 | 1.0000 | 0.18% |
| Arterial Blood Pressure Alarm - High | 0.9964 | 0.9999 | 0.12% |
| Total Bilirubin | 0.9966 | 1.0000 | 0.07% |
| O2 Saturation Pulseoxymetry Alarm - High | 0.9967 | 1.0000 | 0.36% |
| Hematocrit (serum) | 0.9968 | 1.0000 | 0.26% |
| Lactic Acid | 0.9968 | 1.0000 | 0.14% |
| Heart Rate Alarm - Low | 0.9968 | 1.0000 | 0.36% |
| Non-Invasive Blood Pressure Alarm - Low | 0.9969 | 1.0001 | 0.27% |
| Platelet Count | 0.9971 | 1.0001 | 0.23% |
| BUN | 0.9973 | 1.0000 | 0.25% |
| Creatinine (serum) | 0.9975 | 1.0000 | 0.25% |
| Glucose (serum) | 0.9975 | 1.0001 | 0.25% |
| Chloride (serum) | 0.9976 | 1.0000 | 0.26% |
| Prothrombin time | 0.9981 | 1.0000 | 0.16% |
| Resp Alarm - Low | 0.9989 | 1.0019 | 0.35% |
