## Supplementary material for "Continuous Value Tokenization Improves Medical Event Foundation Models": TRIPOD AI Checklist

**Study type:** Development (D) — comparison of three tokenization strategies for a medical event foundation model

**Reference:** Collins GS, Moons KGM, Dhiman P, et al. TRIPOD+AI statement: updated guidance for reporting clinical prediction models that use regression or machine learning methods. BMJ 2024;385:e078378. doi:10.1136/bmj-2023-078378

| Section/Topic | Item | D/E | Checklist Item | Page |
|---|---|---|---|---|
| **TITLE** | | | | |
| Title | 1 | D;E | Identify the study as developing or evaluating a multivariable prediction model, the target population, and outcome | 1 |
| **ABSTRACT** | | | | |
| Abstract | 2 | D;E | See TRIPOD+AI for Abstracts checklist | 1–2 |
| **INTRODUCTION** | | | | |
| Background | 3a | D;E | Explain the healthcare context and rationale for the prediction model, including references to existing models | 3 |
| | 3b | D;E | Describe the target population and intended purpose in the context of the care pathway | 3 |
| | 3c | D;E | Describe any known health inequalities between sociodemographic groups | 20–21 |
| Objectives | 4 | D;E | Specify study objectives, including whether development or validation | 3–4 |
| **METHODS** | | | | |
| Data | 5a | D;E | Describe data sources, rationale, and representativeness | 4–5 |
| | 5b | D;E | Specify dates of collected participant data | 4 |

| Section/Topic | Item | D/E | Checklist Item | Page |
|---|---|---|---|---|
| Participants | 6a | D;E | Specify key elements of the study setting | 4 |
| | 6b | D;E | Describe eligibility criteria | 5 |
| | 6c | D;E | Give details of treatments received and handling | N/A |
| Data preparation | 7 | D;E | Describe data pre-processing and quality checking | 5 |
| Outcome | 8a | D;E | Define the outcome, time horizon, and assessment | 8–10 |
| | 8b | D;E | Describe qualifications of outcome assessors if subjective | N/A |
| | 8c | D;E | Report actions to blind outcome assessment | N/A |
| Predictors | 9a | D | Describe choice and pre-selection of predictors | 5–7 |
| | 9b | D;E | Define all predictors, including how and when measured | 5–7, Tables 1–2, S1 Table |
| | 9c | D;E | Describe qualifications of predictor assessors if subjective | N/A |
| Sample size | 10 | D;E | Explain and justify the study size | 5, 8 |
| Missing data | 11 | D;E | Describe how missing data were handled | 5 |
| Analytical methods | 12a | D | Describe how data were used, including partitioning | 5 |
| | 12b | D | Describe how predictors were handled (transformation, scaling) | 6–7 |
| | 12c | D | Specify model type, rationale, building steps, hyperparameter tuning, internal validation | 7–8 |

| Section/Topic | Item | D/E | Checklist Item | Page |
|---|---|---|---|---|
| | 12d | D;E | Describe handling of heterogeneity across clusters | N/A |
| | 12e | D;E | Specify all performance measures and plots with rationale | 8–10 |
| | 12f | E | Describe any model updating from evaluation | N/A |
| | 12g | E | Describe how model predictions were calculated | 8–10 |
| Class imbalance | 13 | D;E | State if and how class imbalance was addressed | 9 |
| Fairness | 14 | D;E | Describe approaches to address model fairness | 20–21 |
| Model output | 15 | D | Specify model output, classification details, thresholds | 8–10 |
| Training vs evaluation | 16 | D;E | Identify differences between development and evaluation data | 5 |
| Ethical approval | 17 | D;E | Name IRB/ethics committee and describe consent | 4 |
| **OPEN SCIENCE** | | | | |
| Funding | 18a | D;E | Give funding source and role of funders | Submission system |
| Conflicts of interest | 18b | D;E | Declare conflicts and financial disclosures | 23 |
| Protocol | 18c | D;E | Indicate where protocol can be accessed | Not prepared |
| Registration | 18d | D;E | Provide registration information | Not registered |
| Data sharing | 18e | D;E | Provide details of data availability | 22 |
| Code sharing | 18f | D;E | Provide details of code availability | 22 |
| **PATIENT & PUBLIC INVOLVEMENT** | | | | |

| Section/Topic | Item | D/E | Checklist Item | Page |
|---|---|---|---|---|
| PPI | 19 | D;E | Provide details of patient and public involvement | 4 |
| **RESULTS** | | | | |
| Participants | 20a | D;E | Describe flow of participants, with and without outcome | 10–12 |
| | 20b | D;E | Report characteristics overall and per data source | 10–12, Tables 1–2 |
| | 20c | E | Compare predictor distributions with development data | N/A |
| Model development | 21 | D;E | Specify number of participants and outcome events | 10–16 |
| Model specification | 22 | D | Provide full model details for replication | 7–8 |
| Model performance | 23a | D;E | Report performance with CIs, including subgroups | 11–18, Tables 3–4 |
| | 23b | D;E | Report heterogeneity in performance across clusters | N/A |
| Model updating | 24 | E | Report results from any model updating | N/A |
| **DISCUSSION** | | | | |
| Interpretation | 25 | D;E | Interpret main results including fairness, in context of objectives and prior studies | 19–21 |
| Limitations | 26 | D;E | Discuss limitations and effects on bias, uncertainty, generalizability | 20–21 |
| Usability | 27a | D | Describe handling of poor quality or unavailable input data | 5 |
| | 27b | D | Specify user interaction requirements and expertise level | 3–4 |
| | 27c | D;E | Discuss next steps for applicability and generalizability | 21 |
